# FRISTS: Interpretable Time Series-Based Heart Failure Risk Prediction

**DOI:** 10.1101/2023.12.20.23299867

**Authors:** Sophia Lin, Xinyu Dong, Fusheng Wang

## Abstract

Heart failure is an incurable medical condition that affects millions of people globally. Developing prediction models is crucial to prevent patients from progressing to heart failure. Current heart failure prediction models struggle with achieving interpretability, a mandatory trait that allows the model to be applied in healthcare, while possessing high real-world accuracy. We introduce FRISTS, a novel HF prediction approach leveraging sequential times series-based modeling, such as long short-term memory (LSTM) networks, and feature selection. Our study utilized an extensive electronic health record (EHR) database and found that FRISTS outperformed traditional AI models (e.g., Random Forest Classifier, Logistic Regression, and Decision Tree) and more complex machine learning techniques (e.g., XGBoost, LSTM), yielding an average F1 score of 0.805 and receiver operating characteristic area under the curve (ROC AUC) value of 0.990. Our SHAP-inspired permutation method enables interpretation of the feature ranking, enhancing the result transparency and paving the way for a new class of interpretable models. This approach holds promise in enhancing clinical decision-making and patient care in the context of heart failure prevention.

## 1 Introduction

An estimated 6.7 million Americans suffer from heart failure (HF), and the five-year HF mortality rate is 52.6% [1]. As HF is incurable but preventable, the development of prediction models is crucial to help patients with high risk of HF. Machine learning (ML) methods have become popular as predictive models supporting clinical decision-making. However, the application of advanced ML models in the medical and healthcare domain has been limited as increases in a model’s complexity generally led to increased performance but decreased interpretability. This is important as physicians need to understand a model’s reasoning in order to use it in the high-risk environment of healthcare. Model interpretability is vital to assist clinical decisions, combat noisy data, and enable human-in-the-loop validation. Deep learning methods provide high accuracy for specific training datasets but are difficult to interpret and drop in accuracy with real-world data.

Electronic health records (EHR) have been adopted in the US and other countries in the past decade. Because of the comprehensiveness of EHR, which offers longitudinal data on the real-world patient population, interest in using it in HF prediction methods has risen. The large-scale use of EHR strengthens the power of the prediction models and allows the model to be easily adapted.

This work proposes Feature Ranked Interpretable Sequential Time Series (FRISTS) a novel ensemble approach that first uses a feature ranking model, e.g., Random Forest Classifier, to transform input data into a more learnable format for a sequential model, e.g., LSTM (Long Short-Term Memory). FRISTS is shown to achieve high prediction accuracy and model interpretability, which is achieved by a SHAP-inspired permutation method to calculate the contribution score of features. Our study considers a patient’s medical history and uses prescribed medication, lab tests, and demographic information in addition to diagnoses, the model uses a more comprehensive overview of patients. We trained FRISTS using the Cerner Health Facts© EHR database that contains nearly 50 million patients across over 600 hospitals with more than 150,000 HF cases.

## 2 Background and Related Work

There are no standard baselines in HF prediction [2]. Panahiazar et al. have shown that utilizing EHR on the Seattle Heart Failure Model (SHFM) improves accuracy [2]. Boosting and support vector machines (SVM) have been used for six-month HF prediction with poor performance [3].

Common features include diagnoses (comorbidities), demographics (age, sex, race, and ethnicity), medications, and lab tests. A study evaluating the RNN model REverse Time AttentIoN model (RETAIN) over Cerner Health Facts© EMR shows that using all four features provides the best result, with an average AUC of 0.823 [4].

Combining LSTM with other models has been used in opioid overdose risk and blood pressure prediction [5][6], which allows the new architecture to consider the temporal effects of disease progression. Random forest models have previously been shown to achieve the best recall and deep learning models achieve the best precision in prediction models using EHR [7]. Feature ranking is a common method of providing model interpretability [6][8].

## 3 Methods

### 3.1 Dataset and pre-processing

This study uses data spanning from 2014 to 2018 from the Cerner Health Facts© EHR database. Health Facts© encompasses a diverse range of medical information including diagnoses through International Classification of Diseases codes (both ICD-9 and ICD-10 and derivatives), medical information through national drug codes (NDCs), lab tests through Healthcare Common Procedure Coding System (HCPCS) codes with categorical indicators and numerical results, and demographic information (age, race, and gender).

HF diagnosis was determined by the receipt of a diagnosis code beginning with “428” or “I50,” the ICD-9 and ICD-10 diagnosis codes for HF. HF patients and non-HF patients were processed differently. A temporal window of up to five encounters was created for each patient, with the encounters being the last five encounters before the first HF diagnosis for HF patients, and the last six excluding the last encounter for each non-HF patient. See Table 1.

**Table 1:**
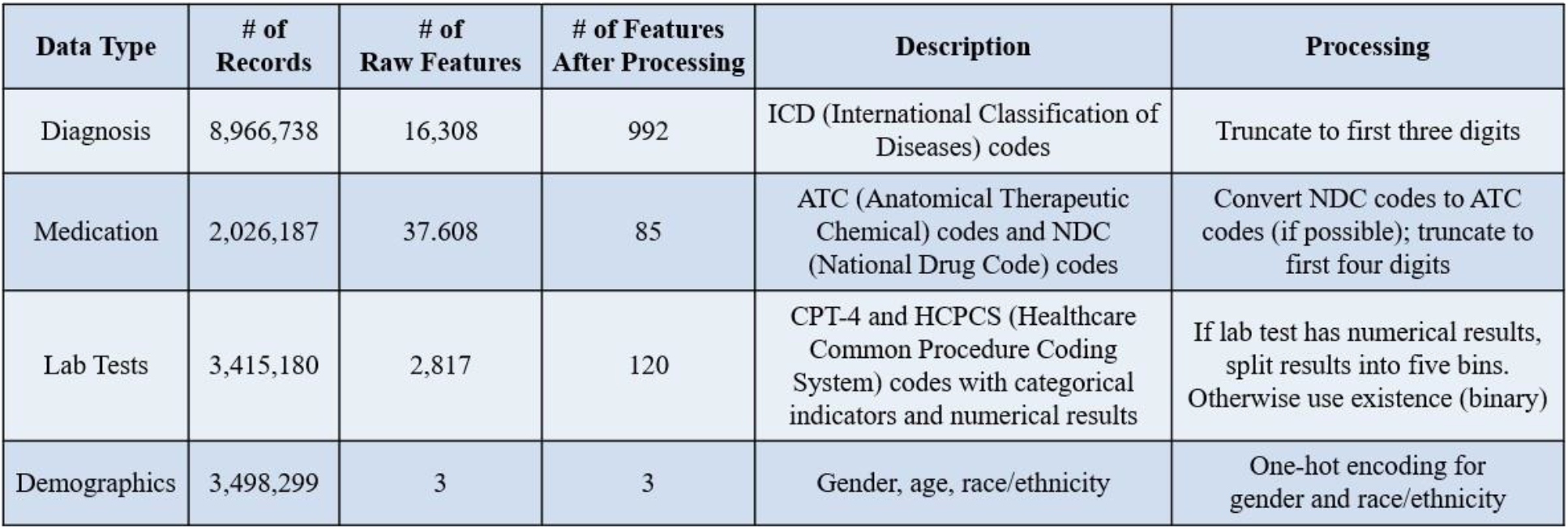
Breakdown of Cerner Health Facts© data.

ICD-9 codes were converted to corresponding ICD-10 codes if they existed. Diagnosis codes were truncated to the first three digits. This groups similar diagnosis codes together and lowers the total number of features. NDC medication codes were converted to corresponding Anatomical Therapeutic Chemical (ATC) code(s) whenever possible. While NDC codes primarily pertain to labeler or manufacturer information, ATC codes are preferred as they focus on the therapeutic attributes and chemical structure of the drug. All medication codes were then truncated to the first four characters. Lab tests were matched to categorical results in the database and then matched to indicators. To reduce bias, we filtered out diagnosis codes, medication codes, and lab tests corresponding to less than 1% of the relevant encounters (i.e., encounters in a patient’s temporal window) in HF patients. This significantly lowered the number of features for consideration (Table 1).

Patients under the age of 16 were removed from the database. The median age of the rest of the dataset was found. For each encounter, the corresponding age measured at the encounter was used when possible. If not, the last measured age before the encounter was used. Patients with no information on age are given the median age. One-hot encoding was applied to race and gender information. We additionally refined race categories to eliminate redundancy (e.g., combining “Asian” and “Asian American”) and address vagueness (e.g., removing “Null” and “Other” categories).

A two-dimensional matrix was created for every patient, with rows representing encounters and columns representing the features (Figure 1). All matrices combine to form a three-dimensional matrix, which was re-shaped as needed for different models.

**Figure 1:**
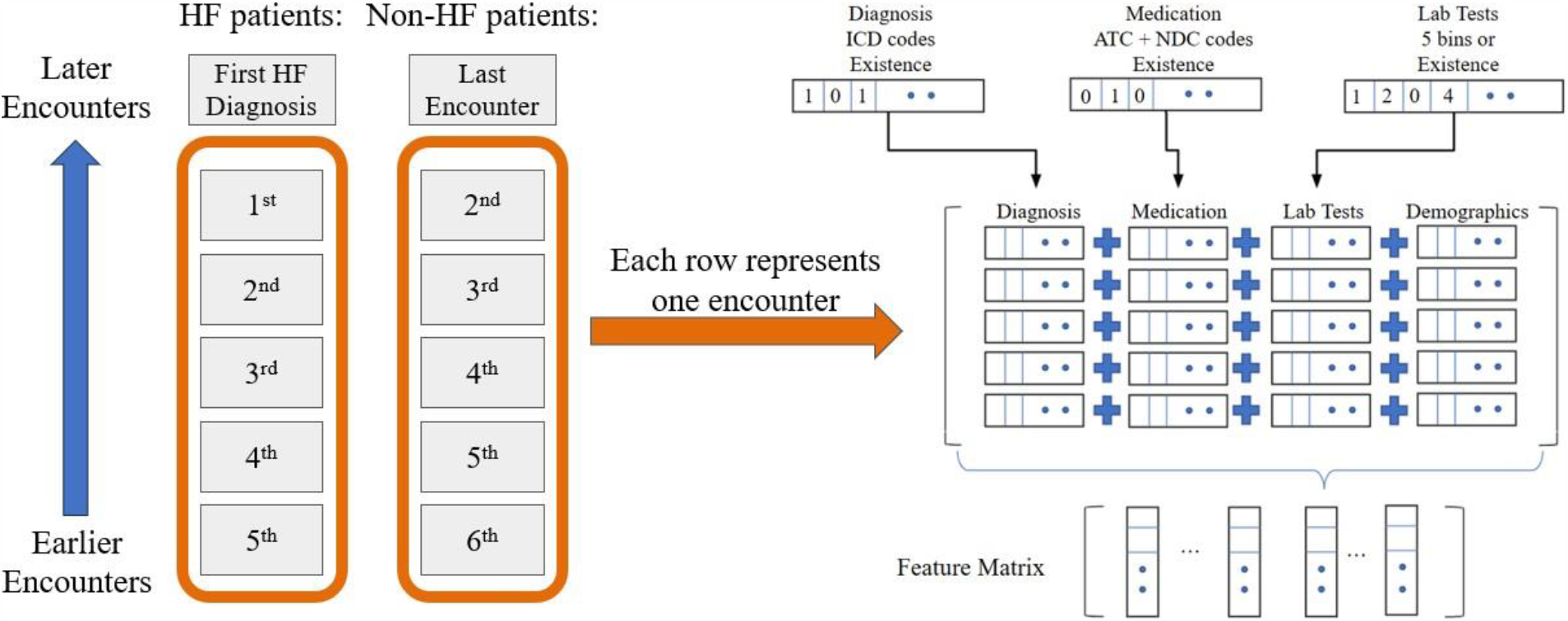
Creation of matrices.

### 3.2 Data processing

The dataset was split into training and testing subsets with an 8:2 ratio.

FRISTS is a multistage approach that first trains a Random Forest Classifier and filters out data features with low importance based on the trained classifier as shown in Figure 2. An LSTM model was then trained on this filtered data using 64 units and 25 epochs. We conducted rigorous benchmarking of FRISTS against the performance of several traditional baselines in the literature (Random Forest Classifier, Decision Tree, Logistic Regression) and advanced techniques such as XGBoost and LSTM.

**Figure 2:**
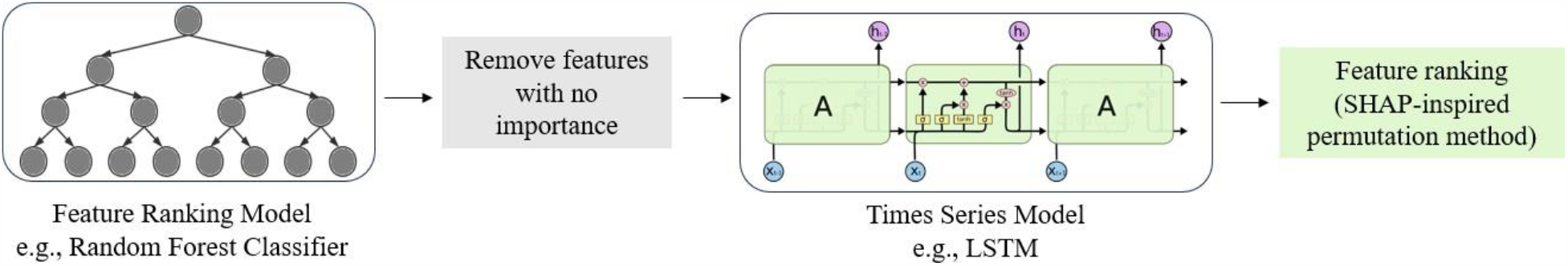
Structure of FRISTS.

### 3.3 Feature ranking

A SHAP-inspired permutation method calculates the contribution score of every feature and produces a feature ranking and learned weights, achieving clinical interpretability. FRISTS’s feature ranking is compared to those of a Random Forest Classifier in Table 4.

## 4 Results

FRISTS outperforms all baseline methods in *F*_1_ and ROC AUC score with an average *F*_1_ score of 0.8045 and ROC-AUC score of 0.9895 (Table 2). The difference in *F*_1_ scores is illustrated in Figure 3.

**Table 2:** *F*_1_ and ROC AUC scores.

| Method Type | Method | Average $F_1$ | Average ROC-AUC |
| --- | --- | --- | --- |
| Traditional Models | Logistic Regression | $0.2311 \pm 0.0141$ | $0.8153 \pm 0.0089$ |
| | Random Forest | $0.2179 \pm 0.0022$ | $0.8142 \pm 0.0040$ |
| | Decision Tree | $0.1201 \pm 0.0077$ | $0.5488 \pm 0.0028$ |
| Boosting | XGBoost | $0.2508 \pm 0.0119$ | $0.8441 \pm 0.0031$ |
| LSTM-based | LSTM | $0.7879 \pm 0.0115$ | $0.9892 \pm 0.0011$ |
| **Ours** | **FRISTS** | **$0.8045 \pm 0.0127$** | **$0.9895 \pm 0.0017$** |

**Figure 3:**
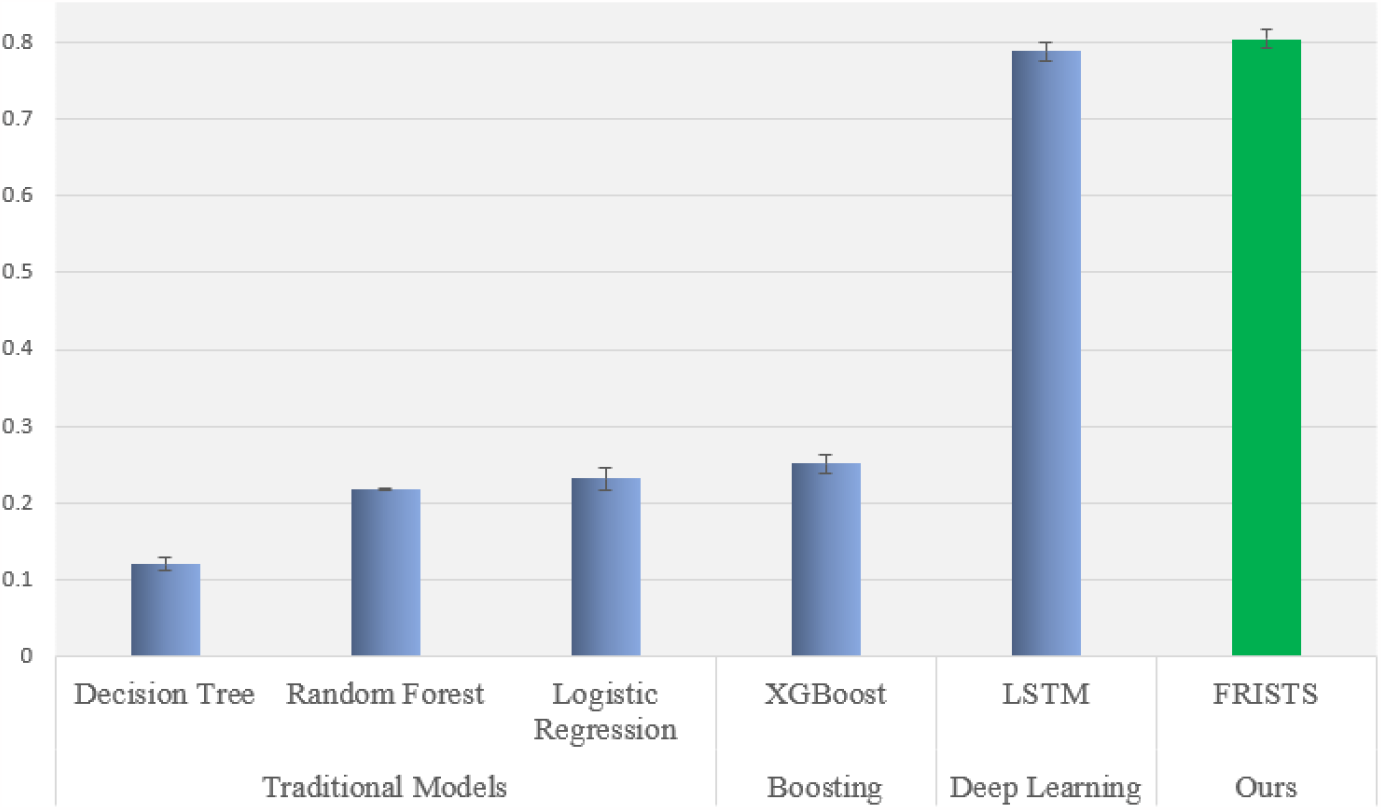
*F*_1_ score comparison. Error ranges are the standard deviation.

The top twenty highest-ranked features for FRISTS are given below in Table 3.

**Table 3:** Twenty highest-ranked features for FRISTS.

| Rank | Category | Description | Rank | Category | Description |
| --- | --- | --- | --- | --- | --- |
| 1 | Demographic | Age | 11 | Medication | GlucaGen (low blood sugar) |
| 2 | Demographic | African American | 12 | Diagnosis | Cardiomyopathy |
| 3 | Diagnosis | Chronic kidney disease | 13 | Diagnosis | Atrial fibrillation and flutter |
| 4 | Diagnosis | Abdominal and pelvic pain | 14 | Diagnosis | Dyspnea, unspecified |
| 5 | Diagnosis | Other chronic obstructive pulmonary disease | 15 | Diagnosis | Other joint disorder, not elsewhere classified |
| 6 | Diagnosis | Encounter for screening for malignant neoplasms | 16 | Diagnosis | Coronary atherosclerosis of native coronary artery |
| 7 | Diagnosis | Diabetes mellitus without mention of complication | 17 | Diagnosis | Complications and ill-defined descriptions of heart disease |
| 8 | Diagnosis | Chronic Ischemic Heart Disease | 18 | Medication | Tylenol Caplet |
| 9 | Diagnosis | Type 2 diabetes mellitus | 19 | Diagnosis | Essential (Primary) Hypertension |
| 10 | Diagnosis | Dilated cardiomyopathy | 20 | Medication | Bupropion Hydrochloride (anti-depression) |

FRISTS’s top-ten feature ranking is compared to that of a Random Forest Classifier (Table 4).

**Table 4:** Comparing the top ten features for FRISTS and a Random Forest Classifier.

| FRISTS |  | Random Forest Classifier |  |
| --- | --- | --- | --- |
| Rank | Description | Rank | Description |
| 1 | Age | 1 | Age |
| 2 | African American | 2 | Symptoms involving respiratory system and other chest symptoms |
| 3 | Chronic kidney disease | 3 | Caucasian |
| 4 | Abdominal and pelvic pain | 4 | Chronic renal failure |
| 5 | Other chronic obstructive pulmonary disease | 5 | Long term (current) drug therapy |
| 6 | Encounter for screening for malignant neoplasms | 6 | African American |
| 7 | Diabetes mellitus without mention of complications | 7 | Occupant of pick-up truck or van injured in non-collision transport accident |
| 8 | Chronic Ischemic Heart Disease | 8 | Type 2 diabetes mellitus |
| 9 | Type 2 diabetes mellitus | 9 | Non-pressure chronic ulcer of left heel and midfoot with muscle involvement without evidence of necrosis |
| 10 | Dilated cardiomyopathy | 10 | Car occupant injured in collision with railway train or railway vehicle |

## 5 Discussions

We found that FRISTS has a nearly four-fold improvement in *F*_1_ compared to traditional baselines, including over a six-fold increase for Decision Tree, as well as over a three-fold increase from the boosting ML model XGBoost. FRISTS even outperforms the deep-learning architecture LSTM in both *F*_1_ and ROC AUC; curiously, FRISTS took a quarter of the training time as LSTM did.

On top of achieving performance gains in numerical results, FRISTS also outputs more interpretable feature rankings that are clearly more relevant to HF than the Random Forest Classifier does (Table 4).

## 6 Conclusion

FRISTS demonstrates remarkable improvement in the accuracy of HF risk prediction while enhancing model interpretability. This novel approach holds promise in enhancing clinical decision-making and patient care in the context of early intervention of HF and can be extended to other model architectures, applications, and datasets.

## Data Availability

All data used in the present study are credential data.

## Acknowledgments

This research was made possible through the support of the Computer Science and Informatics Summer Research Experience (CSIRE) program at Stony Brook University.

## Notes

### Competing Interest Statement

The authors have declared no competing interest.

### Funding Statement

This study did not receive any funding

